# Comparative analysis of variation in the quality and completeness of local outbreak control plans for SARS-CoV-2 in English local authorities

**DOI:** 10.1101/2020.11.30.20240739

**Authors:** Xinming Yu, Mengye Li, Laurie Lawson-Portuphy, Avirup Chowdhury, Padmanabhan Badrinath

## Abstract

**Background:** Local outbreak control plans (LOCPs) are statutory documents produced by local authorities (LAs) across England. LOCPs outlines LAs’ response to COVID-19 outbreaks and the coordination of local resources, data, and communication to support outbreak response. LOCPs are therefore crucial in the nation’s response to COVID-19. However, there has been no previous systematic assessment of these documents. We performed this study to systematically assess the quality of LOCPs and offer recommendations of good practice.

**Methods:** All published LOCPs were assessed for basic characteristics. A framework based on Department of Health and Social Care guidelines was used to assess a random sample of LOCPs. Qualitative analysis was undertaken for LOCPs with highest completeness.

**Results:** 137 of 150 LAs publicly published a full LOCP; nine were drafts. Statistical analysis demonstrated significant difference between reporting of mainstream schools, care homes, and the homeless population and other educational settings, high-risk settings, and other vulnerable groups. LOCPs varied in approach when structuring outbreak response information and focussed on different areas of outbreak management.

**Conclusions:** The majority of LAs are publicly accessible. There is significant variation between the reporting of high-risk settings and groups. Suggested recommendations may help to improve future LOCP updates.

## Introduction

COVID-19, a disease caused by the virus SARS-CoV-2, has serious individual morbidity and mortality, especially among vulnerable populations in our communities (1). The global pandemic has had major economic and social impacts worldwide (2–4), some partly due to restrictions such as lockdown measures. The mental health burden of COVID-19 is further exacerbated by the need for high-risk individuals and the need for people who contracted the virus to self-isolate (4). Care home residents and Black Asian Minority Ethnic (BAME) individuals are disproportionately affected (5–7).

Although COVID-19 is a global pandemic, early identification and response to COVID-19 outbreaks requires an approach tailored to local circumstance (8). The government announced that local authorities (LAs) were to be empowered to take greater responsibility in responding to local COVID-19 outbreaks and to produce a local outbreak control plan (LOCP). LOCPs are central documents, providing information on responding to COVID-19 outbreaks in the local settings, whilst coordinating testing, data use, and communication to support the response. The other key focus of LOCPs is the coordination of resources and support for vulnerable groups and settings, for example supporting self-isolating residents. As England comes out of the national lockdown in December, the country will be placed in the enhanced tiered system (9) and LAs will be receiving dedicated funding (10) for COVID-19 related Public Health activities. Furthermore Christmas COVID-19 guidance (11) has been published. Hence going forward LOCPs will be key to COVID-19 response at a local level, providing the overarching framework for the LA approach and actions.

The government announced that all county councils (CCs), metropolitan districts, unitary authorities, and London boroughs are required to produce and publish LOCPs (12,13). Lower tier LAs were exempt from publishing LOCPs. In order to produce these plans, the government has published a set of guidelines, outlining the focus areas (12). Characteristics of LOCPs that the authors considered important were also incorporated in the assessment.

In this comparative study, we analyse COVID-19 LOCPs in all English LAs. Based on our findings, we provide recommendations for good practices when updating LOCPs.

## Methods

LOCPs from all English LAs were included in the initial analysis of LOCP characteristics. LOCPs were downloaded on the 25^th^ of September 2020 for the unitary authorities and London boroughs. The LOCPs for CCs and metropolitan districts were downloaded on the 7^th^ and 19^th^ of October, respectively.

Random stratified sampling was used to produce a representative sample of LAs for more detailed analysis. The number of LAs were selected to reflect the frequency of their occurrence across the country (14). LAs were stratified by type (CCs, unitary authorities, metropolitan districts, London boroughs) and then by population size (above or below median population size). Unitary authorities were further stratified according to rural-urban classification (15). Metropolitan districts and London boroughs were not stratified this way due to the predominantly urban nature of these districts. Rural-urban stratification was not applicable to CCs. Sampling was performed using random number generation using RStudio (RStudio Inc, USA; version 4.0.2) (16).

The Department of Health and Social Care (DHSC) guidance identifies seven major themes for inclusion in LOCPS: healthcare and education settings; high-risk workplaces, communities and settings; local testing; contact tracing; data integration; vulnerable people and diverse communities; and local boards (12). Data was extracted using a template designed using DHSC guidance and other points the authors considered as important to assess the quality and completeness of LOCPs.

Data extraction and quality assessment were performed by four authors (X.Y., M.L., A.C., and L.L.). If LAs only made publicly available, the summaries of LOCPs then those documents were used. Full LOCPs were used for more in-depth analysis in our sub-sample. Publicly available was defined as being able to find the LOCP on the first page of the search engine or being present on a LAs’ homepage or COVID-19 response webpage.

Data were analysed in RStudio using the *Stringr* (17) packages. We compared well-reported settings (those reported in at least 70% of sampled LOCPs) with those less well reported. The lowest value of well-reported settings was compared with highest value of other settings. Fisher’s exact test was performed, with Bonferroni correction, to check for variation in the reporting of different high-risk settings and vulnerable groups. New significance threshold after correction was (p=0.006). Statistical analysis was performed on GraphPad Prism (GraphPad Prism version 9.0.0 for Mac, GraphPad Software, San Diego, California, USA) (18).

LAs with the highest proportion of fulfilled criteria per setting were taken forward for qualitative analysis in order to maximise the data available for thematic analysis. Where multiple LOCPs had the same number of fulfilled criteria, an LOCP was selected using random number generation in RStudio. Areas examined in qualitative analysis included risk assessment, prevention strategies, testing, supportive measures, communication strategies, and whether the outbreak response plan for settings had a single point of contact (SPOC). The nature of the content (e.g. operational or strategic) was also noted.

## Results

150 LAs published LOCPs; key characteristics are summarised in Table 1. 137 LAs (91%) had a publicly available full LOCP, of which 128 were final documents. Nine published LOCPs were draft versions. Two LAs did not have publicly available LOCPs.

**Table 1.** Key characteristics of published local outbreak control plans (LOCPs), overall and by local authority (LA) subgroup.

|  | Full LOCP publicly available (%) | LOCP easily accessible (%) | Summary publicly available (%) | Finalised LOCP (%) |
| --- | --- | --- | --- | --- |
| **Population** |  |  |  |  |
| **All LAs (n= 150)** | **137 (91%)** | **140 (93%)** | **60 (40%)** | **128 (91%)** |
| **LA by type** |  |  |  |  |
| County councils (n= 26) | 24 (92%) | 26 (100%) | 17 (65%) | 24 (92%) |
| Metropolitan districts (n= 36) | 33 (92%) | 35 (97%) | 11 (31%) | 30 (86%) |
| London boroughs (n= 32) | 32 (100%) | 32 (100%) | 9 (28%) | 27 (84%) |
| Unitary authorities (n= 56) | 48 (86%) | 47 (84%) | 23 (41%) | 47 (100%) |
| *Unitary urban (n= 43)* | 37 (86%) | 36 (84%) | 19 (44%) | 36 (97%) |
| *Unitary rural (n= 13)* | 11 (85%) | 11 (85%) | 4 (31%) | 11 (100%) |
| **Population quartile** |  |  |  |  |
| 1st quartile (n= 38) | 34 (89%) | 34 (89%) | 14 (37%) | 32 (94%) |
| 2nd quartile (n= 37) | 35 (95%) | 35 (95%) | 12 (32%) | 30 (86%) |
| 3rd quartile (n= 37) | 32 (86%) | 34 (92%) | 14 (38%) | 30 (88%) |
| 4th quartile (n= 38) | 36 (95%) | 37 (97%) | 20 (53%) | 36 (95%) |

140 (93%) LOCPs or summary documents were easily accessible, according to the criteria listed in methods. 60 (40%) of LAs had a LOCP summary document publicly available, either accompanying a full LOCP or on their own. CCs had the longest full LOCPs, at an average of 53 pages. Unitary authorities and metropolitan districts had an average length of 41 and 37 pages respectively. London LOCPs were on average 34 pages long. 23 (16%) of all LOCP and summaries had a review date.

A sample of 30 LOCPs was selected for further analysis: 6 from CCs, metropolitan districts, and London boroughs, and 12 from unitary authorities. All sampled LOCPs were full documents.

93% of LOCPs outlined local testing infrastructure. 80% of LOCPs have identified local testing facilities, including laboratories and mobile testing units. 77% of LOCPs have mentioned or included the coordination of local, regional, and national testing capabilities. 63% of LOCPs considered or outlined measures for increasing test capacity in response to increasing pressures. 77% of LOCPs made reference to directing and deploying testing for contact tracing.

Data integration for supporting outbreak management was mentioned in 63% of LOCPs. Integrating data to support contact tracing (37%), and monitoring of effectiveness and impact (53%) were mentioned in a proportion of sampled LOCPs.

Governance was covered by the majority of LAs. 80% of LOCPs outlined which Boards were involved in the outbreak response and how the Boards communicate. 63% of LOCPs outlined the memberships of each Board.

Overall and for unitary authorities, the City of York Council’s LOCP had fulfilled the most criteria of our template. This was also the case for Croydon, North Tyneside Borough Council, and Nottinghamshire for London boroughs, metropolitan districts, and CC respectively.

Table 2 outlines reporting variation across healthcare and education, care and vulnerable groups; certain groups were more likely to be reported than others. Special education settings (20%) and boarding schools (10%) were reported significantly less often than early years schools (73%) (p=0.0001); extra care housing (10%) and support housing reporting (23%) were lower than for residential nursing homes (77%) (p=0.0001); reporting of BAME (23%) and travellers (23%) were significantly lower than the homeless group (70%) (p=0.001); reporting of migrants (20%), and asylum seekers and refugees (20%) were significantly lower than the homeless group (70%) (p=0.0002).

**Table 2.** Proportion of local authorities (LAs) reporting selected high-risk settings in local outbreak control plans. Settings compared to early years schools *(p=0.0001). Settings compared to residential nursing homes ** (p=0.0001). Settings compared to the homeless population ***(p=0.001), †(p=0.0002).

| Population | <u>Health care and education settings</u> |  |  |  |  |  | <u>High-risk workplaces, communities and locations</u> |  |  |  |  | <u>Vulnerable people and diverse communities</u> |  |  |  |  |  |
| --- | --- | --- | --- | --- | --- | --- | --- | --- | --- | --- | --- | --- | --- | --- | --- | --- | --- |
|  | Primary schools | Secondary schools | Early years schools | Special education settings | Boarding schools | Primary health care | Care homes | Nursing homes | Support housing | Extra care housing | Workplaces | Homeless population | Vulnerable people | Black Asian Minority Ethnic | Travellers | Migrants | Asylum seekers and refugees |
| **All sampled LAs (n= 30)** | **24 (80%)** | **22 (73%)** | **22 (73%)** | **6 (20%)\*** | **3 (10%)\*** | **11 (37%)** | **24 (80%)** | **23 (77%)** | **7 (23%)\*\*** | **3 (10%)\*\*** | **21 (70%)** | **21 (70%)** | **11 (37%)** | **7 (23%)\*** | **7 (23%)\*** | **6 (20%)\*** | **6 (20%)\*** |
| **LAs by population size** |  |  |  |  |  |  |  |  |  |  |  |  |  |  |  |  |  |
| ≤ 50th centile (n= 16) | 14 (88%) | 13 (81%) | 15 (94%) | 3 (19%) | 2 (13%) | 7 (44%) | 13 (81%) | 13 (81%) | 4 (25%) | 2 (13%) | 11 (69%) | 12 (75%) | 8 (50%) | 4 (25%) | 3 (19%) | 4 (25%) | 4 (25%) |
| > 50th centile (n= 14) | 10 (71%) | 9 (64%) | 7 (50%) | 3 (21%) | 1 (7%) | 4 (29%) | 11 (79%) | 10 (71%) | 3 (21%) | 1 (7%) | 10 (71%) | 9 (64%) | 3 (21%) | 3 (21%) | 4 (29%) | 2 (14%) | 2 (14%) |
| **LAs by type** |  |  |  |  |  |  |  |  |  |  |  |  |  |  |  |  |  |
| County councils (n= 6) | 5 (83%) | 5 (83%) | 3 (50%) | 2 (33%) | 1 (17%) | 2 (33%) | 5 (83%) | 4 (67%) | 2 (33%) | 1 (17%) | 5 (83%) | 5 (83%) | 1 (17%) | 2 (33%) | 2 (33%) | 1 (17%) | 1 (17%) |
| Metropolitan districts (n= 6) | 4 (67%) | 3 (50%) | 3 (50%) | 2 (33%) | 1 (17%) | 2 (33%) | 4 (67%) | 4 (67%) | 1 (17%) | 0 (0%) | 4 (67%) | 3 (50%) | 2 (33%) | 0 (0%) | 0 (0%) | 1 (17%) | 1 (17%) |
| London boroughs (n= 6) | 5 (83%) | 4 (67%) | 6 (100%) | 0 (0%) | 0 (0%) | 4 (67%) | 4 (67%) | 4 (67%) | 2 (33%) | 2 (33%) | 4 (67%) | 5 (83%) | 1 (17%) | 2 (33%) | 1 (17%) | 1 (17%) | 1 (17%) |
| Unitary authorities (n= 12) | 10 (83%) | 10 (83%) | 10 (83%) | 2 (17%) | 1 (8%) | 3 (25%) | 11 (92%) | 11 (92%) | 2 (17%) | 0 (0%) | 8 (67%) | 8 (67%) | 7 (58%) | 3 (25%) | 4 (33%) | 3 (25%) | 3 (25%) |

Qualitative analysis showed variation in content across key LOCP areas (Table 3). Some LOCPs were more operational in content, containing very specific instructions for responding to outbreaks in specific high-risk settings and vulnerable groups. Other plans were strategic in nature, focussing more on high-level approaches to local outbreak response.

**Table 3.**
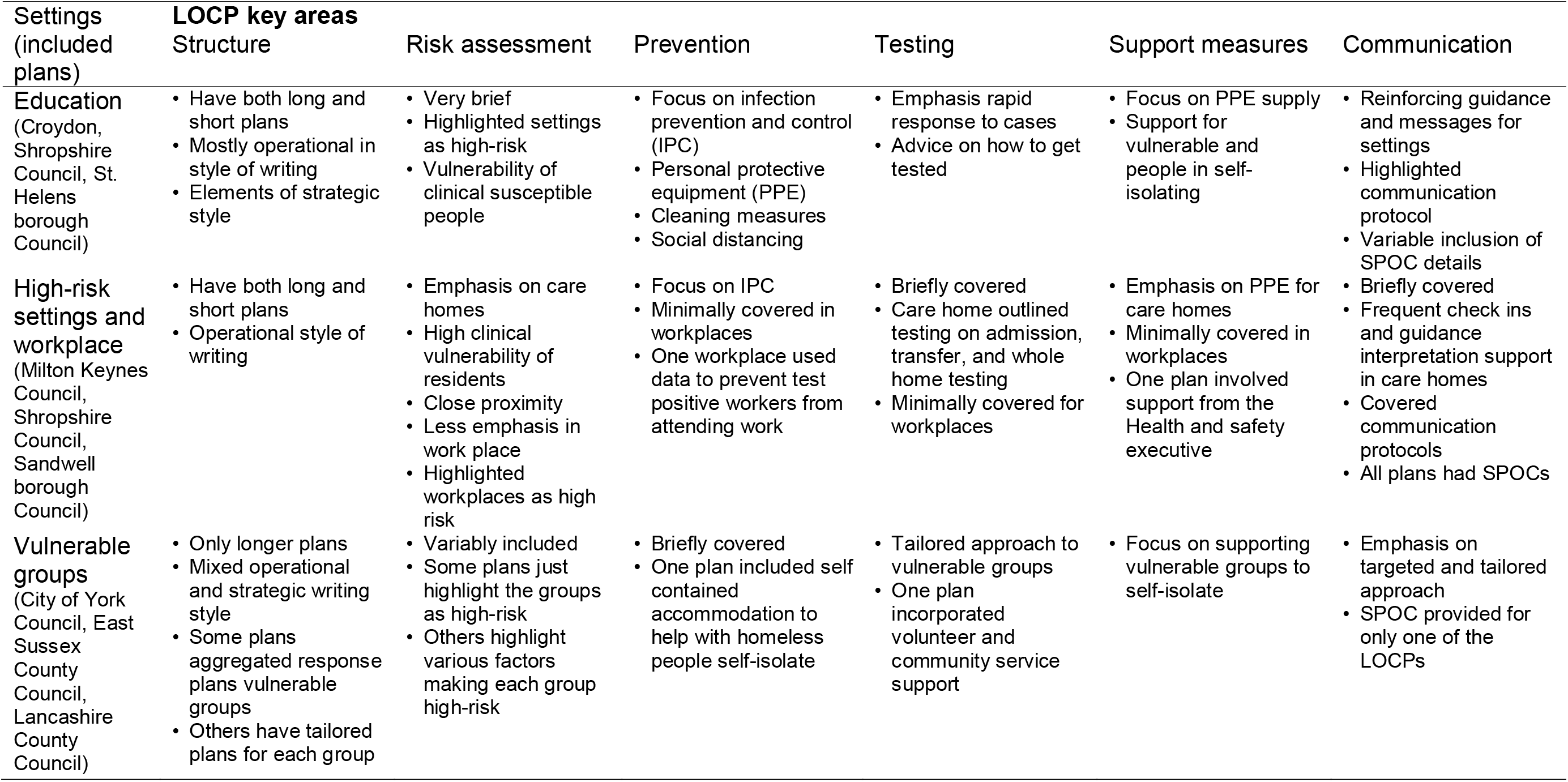
Results of qualitative assessment of well reported settings in complete local outbreak control plans (LOCPs), by high-risk areas.

The contents of outbreak response to each setting were variable. LOCPs had varying levels of risk assessment: some were detailed, for example identifying proximity and clinical vulnerability as risk factors in care homes, where others only categorised settings as high-risk. Prevention strategies had a different focus depending on setting. Common emerging themes included rapid response to cases, advice on getting tested, preventing positive workers from going to work, and support for homeless people to self-isolate. Prevention themes focussed on infection prevention and control and personal protective equipment (PPE). Testing themes covered whole care home testing. Incorporating capabilities from community and volunteering services to support the vulnerable, PPE supply, supporting self-isolating people, and incorporating Health and Safety Executive capabilities were some of the supportive measures identified. Communication support involved interpreting government guidelines, disseminating new information, targeted communication approaches, and outlining the communication protocol in the event of outbreaks.

## Discussion

### Main findings of this study

Government guidance has stated that all eligible LAs are required to produce a LOCP. The majority of LAs across England had published a publicly available LOCP, either as a complete or a summary document. For two LAs, we could not access either documents. This does not exclude the possibility that these are available as internal documents. However, if LOCPs have not been shared, partners and vulnerable settings would lack the ability to access vital information for responding to COVID-19 outbreaks. Examples include contact information for the SPOC or information on available support.

Out of the full LOCPs, nine were in the draft stages. This may be problematic as summary documents do not have as much content as full LOCPs and are less operational; drafts are also subject to major revisions and may have missing information. A notable proportion of LAs (40%) had published LOCP summary documents. These tended to be shorter and may be more suitable as public-facing documents which give those unfamiliar with LOCPs an overview of contents. LAs should consider producing summary LOCPs if they have not already done so.

The average length of full LOCPs varied across LA types and could be interpreted as a surrogate measure for the amount of content included in the plan. The longer length of CC plans may reflect a need to be more detailed, and the need to cover more settings. CCs may need to be detailed because of the larger geographic region covered. Each London borough covers a relatively smaller geographical area, with adjacent neighbouring boroughs having their own plans. Therefore, London LOCPs could be less detailed because of the high density of LAs producing LOCPs in London. In-depth analysis, however, revealed that the plans do not necessarily have to be longer to cover most assessment criteria. When analysing which LOCPs had the greatest number of fulfilled setting specific assessment criteria, a mix of shorter and longer plans were found.

It may be important for LOCPs to be regularly reviewed to reflect the rapidly changing knowledge base and government policy surrounding COVID-19. A low proportion of all LOCPs had a review date in place, and many of these were overdue at the time of data collection. A structured approach to review dates and methods may ensure that LOCPs are up to date.

Statistical analysis revealed significant variation between reporting of high-risk settings and groups in LOCPs. Mainstream educational settings were found to be well reported across all sampled LAs, while other educational settings, such as special education settings and boarding schools, were less reported in outbreak control plans. A similar trend can be found in high-risk settings and in the vulnerable groups category. Care homes and the homeless were reported significantly more than other settings and vulnerable groups. This difference in reporting could be due to a number of factors. The government has allocated significant funding for outbreak response in homeless settings (19,20), potentially increasing the focus upon this group. Vulnerability of settings and groups may also factor into reporting. Care homes have been an area of major concern during the COVID pandemic (6), with major media and public attention; high reporting in LOCPs may be expected as a consequence. However, increased focus on specific settings may result in neglect of others.

Lack of reporting poses a major issue for local outbreak response. Different settings will have varying requirements and challenges, and some pose specific barriers to controlling outbreaks. For example, social distancing can be an issue in the special education setting (21) and travelling students may pose an issue in boarding schools, due to the need to self-isolate (22). Therefore, a tailored approach is required to effectively respond to COVID outbreaks and so LOCPs should try and incorporate further breadth and consider implementing outbreak response plans for these alternative settings.

The structure and content of key areas in well-reported LOCPs were variable between the different plans. Some LOCPs preferred to organise the response to outbreaks in different settings and groups by discrete sections, while others aggregated different groups and settings, covering requirements by theme. For example, they may organise the plan in terms of prevention, communication, and supportive strategies and then discuss individual settings within each section. Either approach may be suitable to achieve sufficient LOCP breadth and detail.

It was found that LOCPs covering the same theme can have different focus areas. For example, when looking at communication strategies in educational settings, one LOCP may focus on the management of communication during an outbreak, with the media and parent groups, whereas another focusses on rapid communication to disseminate information and changes in guidance. We have also noticed that different settings will have different focus areas: testing and communication tended to be emphasised in education settings, whereas care homes focussed more on supportive measures. This could be due to LOCPs being tailored to local circumstances and needs. LAs will likely have differing priorities and local resources to support outbreak response and so may need to adapt their LOCPs accordingly. However, this may result in certain areas being neglected, reducing the effectiveness of outbreak response.

### What is already known on this topic

To our knowledge there has been no published literature assessing the quality of the LOCPs across the country. This is the first study analysing characteristics and coverage of LOCPs.

### What this study adds

The quality of LOCPs, including how comprehensive and user friendly these documents are, is crucial to bringing the pandemic under control. Although the fight is happening across various fronts, the most important aspect is the fight at the local level led by the Directors of Public Health (DPHs) and their teams. LOCPs are the key battle plans at the local level. It would be of help to the DPHs and the central government to understand factors which constitute a good LOCP so that any limitations can be addressed when the plans are next updated. Our study reviews and compares COVID-19 LOCPs in LAs across England and provides recommendations for good practices when producing COVID-19 LOCPs.

Based on our finds, we make the following recommendations for LAs and others:

1. All LAs should publish a finalised LOCP.
2. A summary document should be published alongside full LOCPs to facilitate public transparency
3. Review dates and regular review mechanisms should be incorporated into LOCP production to ensure they are responsive to rapidly changing COVID-19 guidelines.
4. LOCPs should consider outbreak responses in underrepresented high-risk settings, vulnerable groups, and educational settings.
5. Association of DPHs should run a similar competition on LOCPs as they do for the Annual Public Reports (23).

### Limitations

LAs without publicly available LOCPs may still have LOCPs available within the authority. Likewise, not mentioning criteria within the LOCP does not exclude that the LA from having processes in place. However, given the public scrutiny facing LAs in the COVID-19 era, it may be prudent to make these processes more transparent. This is especially so with more implicit processes and is something that we have found when assessing LOCPs using the Association of Directors of Public Health guidance, which involved mostly implicit processes (24). For example, the guidance recommends that the LAs should have robust commissioning processes when delivering outbreak response functions. We found that LAs were unlikely to explicitly include information like this in LOCPs. Therefore, we were not able to account for the extent to which implicit processes have been accounted for.

The small sample size of each group in each LA type, six and twelve, limited the statistical analyses which could be undertaken. Our method of assessing which plan fulfilled the most criteria during assessment was quantitative in nature. This may bias our results toward LOCPs covering a greater number of areas but with less individual detail.

There was some variation in definitions across high-risk settings, for example the definition of vulnerable people. This may result in differences in interpretation: areas with broader definitions of a given area may appear to be more inclusive for a single setting, whilst appearing to lack breadth.

This study examined only specific high-risk settings; it remains a possibility that there are deficiencies in LOCPs elsewhere. Future studies are needed to characterise other settings in more detail.

## Conclusion

This study compares characteristics of LOCPs published by English LAs. The majority of LAs have published a form of LOCP, although some were in draft form or lacked summary documents. LOCPs varied in length, with London boroughs tending to have shorter plans than other LA types. On detailed analysis of a subset of plans, we identified a statistically significant difference in reporting within high-risk settings. This may reflect the impact of media and public interest, or local priorities. Qualitative analysis identified different approach of structuring LOCPs and different focus themes across high-risk settings. We have identified recommendations which may help to improve future updates of LOCPs.

## Data Availability

The data underlying this article are available in the article and in its online supplementary material.

